## Supplemental File for "Optimizing testing for COVID-19 in India"

January 1, 2021

#### S1 Reproduction Number: Next Generation Matrix Method

We start off by rearranging the equations into two different compartments, infected and non-infected:

$$X = \begin{pmatrix} A \\ P \\ MI \\ SI \\ H \end{pmatrix}, \qquad Y = \begin{pmatrix} S \\ R \end{pmatrix}$$

Defining two new vectors $\mathcal{F}$ and $\mathcal{V}$ such that

$$\frac{\mathrm{d}X}{\mathrm{d}t} = (\mathcal{F} - \mathcal{V}),$$

where $\mathcal{F}$ is the vector of **new** infection rates (flows from $Y$ to $X$), and $\mathcal{V}$ is the vector of all **other** rates (no new infections), which include flows from $X$ to $Y$, and flows within $X$. For each compartment, the inflow terms in $\mathcal{V}$ are negative, and the outflow is positive.

From these vectors, we can define two matrices

$$F = \left(\frac{\partial \mathcal{F}}{\partial X}\right)\bigg|_{\text{At DFE}} \qquad V = \left(\frac{\partial \mathcal{V}}{\partial X}\right)\bigg|_{\text{At DFE}},$$

such that the next generation matrix is $G = FV^{-1}$. The reproduction number $R_0$ is the spectral radius (i.e. maximum eigenvalue) of $G$, i.e. $R_0 = \rho(FV^{-1})$.

**Note:** The Disease Free Equilibrium occurs when $I = A + P + MI + SI + H = 0$, i.e. when $\dot{S} = 0$, or $S = N$.

$$\mathcal{F} = \begin{pmatrix} \gamma \frac{\lambda_S}{N} S(A+P+MI+SI+H) \\ (1-\gamma)\frac{\lambda_S}{N} S(A+P+MI+SI+H) \\ 0 \\ 0 \\ 0 \end{pmatrix}$$

$$\implies F = \frac{\lambda_S}{N} S^* \begin{pmatrix} \gamma & \gamma & \gamma & \gamma & \gamma \\ (1-\gamma) & (1-\gamma) & (1-\gamma) & (1-\gamma) & (1-\gamma) \\ 0 & 0 & 0 & 0 & 0 \\ 0 & 0 & 0 & 0 & 0 \\ 0 & 0 & 0 & 0 & 0 \end{pmatrix}$$

$$\mathcal{V} = \begin{pmatrix} \lambda_A A \\ \lambda_P P \\ \lambda_{MI} MI - \delta \lambda_P P \\ \lambda_{SI} SI - (1-\delta)\lambda_P P \\ \lambda_H H - \sigma \lambda_{SI} SI \end{pmatrix} \implies V = \begin{pmatrix} \lambda_A & 0 & 0 & 0 & 0 \\ 0 & \lambda_P & 0 & 0 & 0 \\ 0 & -\delta\lambda_P & \lambda_{MI} & 0 & 0 \\ 0 & -(1-\delta)\lambda_P & 0 & \lambda_{SI} & 0 \\ 0 & 0 & 0 & -\sigma\lambda_{SI} & \lambda_H \end{pmatrix}$$

Using this we can compute the next-generation matrix,

$$G = FV^{-1} =$$

$$\begin{pmatrix} \frac{\gamma\lambda_S}{\lambda_A} & \gamma\lambda_S\left(\frac{1}{\lambda_P} + \frac{\delta}{\lambda_{MI}} + (1-\delta)\left(\frac{1}{\lambda_{SI}} + \frac{\sigma}{\lambda_H}\right)\right) & \frac{\gamma\lambda_S}{\lambda_{MI}} & \gamma\lambda_S\left(\frac{1}{\lambda_{SI}} + \frac{\sigma}{\lambda_H}\right) & \gamma\frac{\lambda_S}{\lambda_H} \\ \frac{(1-\gamma)\lambda_S}{\lambda_A} & (1-\gamma)\lambda_S\left(\frac{1}{\lambda_P} + \frac{\delta}{\lambda_{MI}} + (1-\delta)\left(\frac{1}{\lambda_{SI}} + \frac{\sigma}{\lambda_H}\right)\right) & \frac{(1-\gamma)\lambda_S}{\lambda_{MI}} & (1-\gamma)\lambda_S\left(\frac{1}{\lambda_{SI}} + \frac{\sigma}{\lambda_H}\right) & \frac{(1-\gamma)\lambda_S}{\lambda_H} \\ 0 & 0 & 0 & 0 & 0 \\ 0 & 0 & 0 & 0 & 0 \\ 0 & 0 & 0 & 0 & 0 \end{pmatrix}$$

The eigenvalues are $\left\{0, 0, 0, 0, \lambda_S\left(\frac{\gamma}{\lambda_A} + (1-\gamma)\left(\frac{1}{\lambda_P} + \frac{\delta}{\lambda_{MI}} + (1-\delta)\left(\frac{1}{\lambda_{SI}} + \frac{\sigma}{\lambda_H}\right)\right)\right)\right\}$. Thus

$$R_0 = \lambda_S\left(\frac{\gamma}{\lambda_A} + (1-\gamma)\left(\frac{1}{\lambda_P} + \frac{\delta}{\lambda_{MI}} + (1-\delta)\left(\frac{1}{\lambda_{SI}} + \frac{\sigma}{\lambda_H}\right)\right)\right).$$

Using the parameters given above:

$$R_0 = 2.374.$$

#### S2 Scaling up our simulation

The results of the main text are obtained for a population size of 10,000 individuals, with 2,750 locations (2,500 home locations and 250 work locations). This corresponds to a Poisson distributed population in the home and work locations with a mean of 4 and 40 individuals respectively. We also started with an initial infection seed of 10 asymptomatic individuals (0.1% of the population). However, we also tried a number of runs that were scaled-up to larger population sizes (while keeping the initial infection fraction the same, and preserving household and work location sizes) in order to check for finite-size effects. The results of these simulations are shown in Fig. 2.1, and they are found to agree remarkably well with our results for 10,000 individuals, indicating that this problem scales well with the total population.

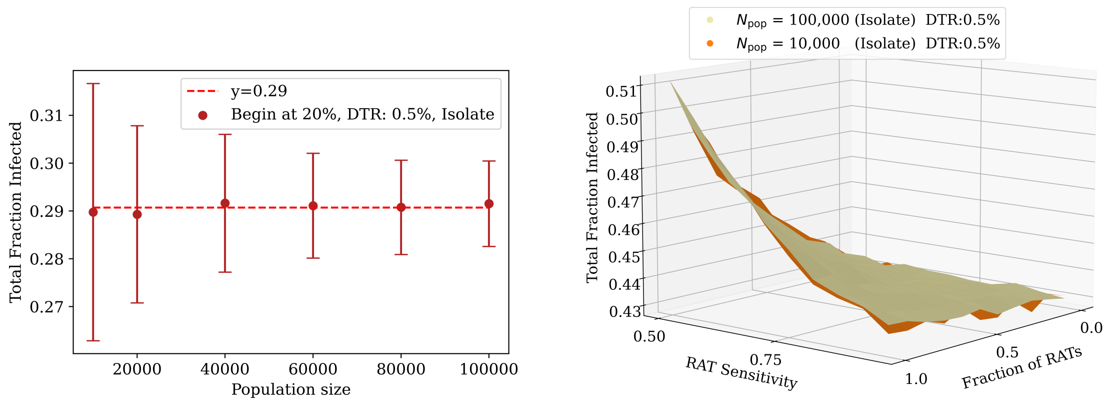


**(a)** Variation with population size. (DTR = 0.5%, starting at 20% recovered, only PCR tests)

**(b)** Stacks for a DTR 0.5%, starting at 10% recovered with $N_{pop} = 10,000$ and $100,000$.

**Fig 2.1: Effects of scaling up the population.** (a) Runs were conducted for different population sizes up to 100,000, with a daily testing rate of 0.5% and testing starting when 20% of the population had recovered. The results of the total infected fraction are shown for each of these populations, with the horizontal line representing their least-square fit. (b) A stack similar to those shown in the main text was also produced to compare the results for population sizes of 10,000 and 100,000, with a daily testing rate of 0.5% and with testing starting at 10%. The results show agreement over the range of RAT test sensitivities and fractions in the mixture. In both the above cases, the individuals who tested positive were isolated when they were declared.

### S3 The effects of Random Testing

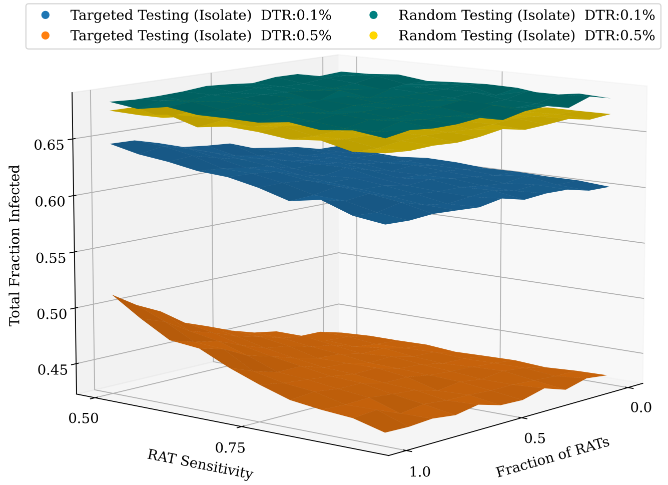


**Fig 3.1: Effects of random testing.** If the testing is assumed to be purely random, with individuals picked out randomly from the population with no preference given to symptomatic individuals, the efficiency of testing is extremely low, as can be seen. While an increase in the daily testing does reduce the total infected fraction at the end of the pandemic, it is nowhere near as effective as targeted testing at the testing rates employed, demonstrating that it is extremely desirable to target symptomatics.

#### S4 Effects of test delays on quarantining strategies

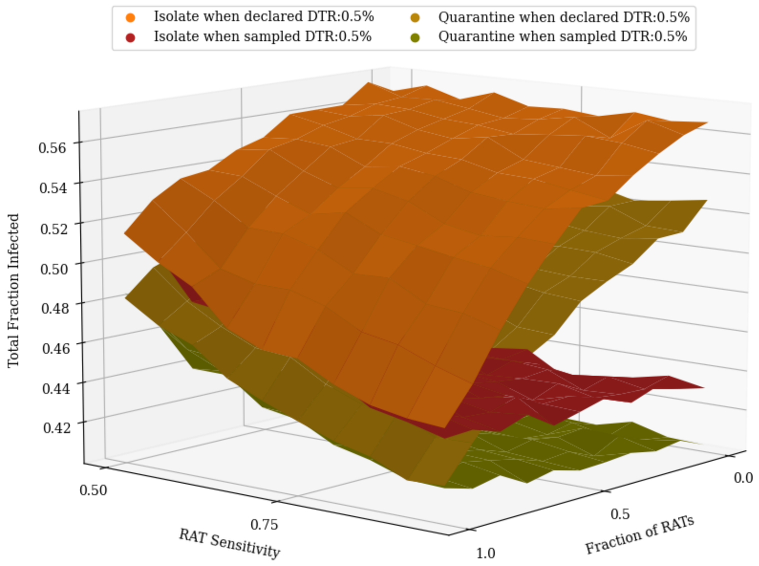


**(a)** Comparing all quarantining strategies for DTR 0.5%

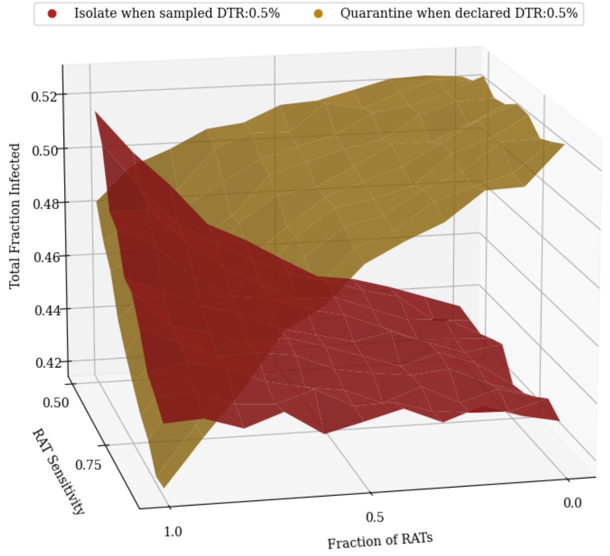


**(b)** Comparing isolating individuals when sampled with quarantining homes when declared

**Fig 4.1: Comparing quarantining strategies with test delays.** Four different quarantining procedures are shown for a daily testing rate of 0.5%. Testing begins when 20% of the population has recovered. PCR tests have a 5 day delay, while RAT remain point-of-care tests. (a) The benefit of having more PCR tests in the mixture can be regained if the interventions are enforced when the test sample is taken. This effect is more significant at higher testing rates, though the trends remain the same. (b) At RAT:PCR ratios of 80:20, quarantining homes of those with positive results can do as well or even better than isolating all test candidates when they are sampled.

When no test delays are included, the results of tests are obtained immediately. There are thus only two different quarantining strategies possible:

(a) **Isolate:** Isolate the individual at home, reducing their infectivity and restricting their movement for 14 days,

(b) **Isolate and quarantine:** In addition to the above, also quarantine the entire home by restricting movement of all family members.

However, with the addition of a test delay, we can now make a distinction between enforcing the intervention when the individual is *sampled* and enforcing it when the result is *declared.* As a result, four different quarantining strategies are now possible:

(a) **Isolate when declared:** Isolate the individual at home, reducing their infectivity and restricting their movement for 14 days when the individual tests positive,

(b) **Isolate and quarantine when declared:** In addition to the above, also quarantine the entire home by restricting movement of all family members when the individual tests positive.

(c) **Isolate when sampled:** Isolate the individual at home, reducing their infectivity and restricting their movement when the individual is sampled for a test. If the test result is negative, the individual is released from isolation. If it is positive, they are further isolated for 14 days.

(d) **Isolate and quarantine when sampled:** In addition to the previous point, the individual's home is also quarantined until the result is declared. If it is negative, the quarantine is lifted, but if it's positive the home remains quarantined.

Fig. 4.1 shows the effect of these different strategies. As shown in the main text, the benefit of the PCR tests' sensitivity is offset by the introduction of the delay, and using only PCR tests is no longer favourable. However, this can be countered by quarantining individuals or homes when the samples are taken. Quarantining homes still makes a larger dent in the total fraction of infected, however when large fractions of RAT tests are used (RAT:PCR $\sim$ 80:20) equivalent or marginally better results can be obtained by quarantining the homes of the tested individual when they are declared positive, instead of isolating them when they are sampled.

### S5 Cost of imposing interventions when test sample is taken

Fig. 5.1 shows the total cost in terms of confining individuals and homes as a function of time, given different quarantining strategies. Strategies that involve interventions being enforced when the individual is sampled are found to require a larger number of people and homes confined. We consider the case when only PCR tests are used with a delay of 5 days between sampling and the results being declared, since this leads to the largest number of people and homes quarantined. In the case of interventions that are enforced when the individual is sampled for a test, the number of people or homes confined does not die out even after the pandemic has passed, since people continue to be tested and remain confined for a duration of 5 days until the PCR result is declared. The peak number of people confined varies from 4% to 6%, and this translates into a fraction of homes quarantined between 15% and 25%.

### S6 Synergy between Testing and Masking

As shown in the main text, there exists a synergy between testing and masking In Fig. 6.1, we compare the effect that masking would have had if the effect had been purely additive or mul-

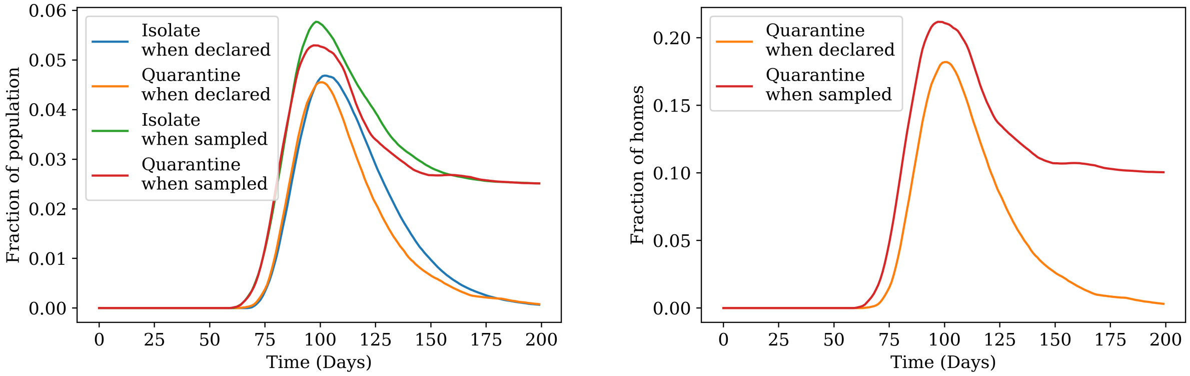


**(a)** Comparing number of individuals isolated **(b)** Comparing number of homes quarantined

**Fig 5.1: People and homes confined as a function of time.** The plot shows the number of people confined on any given day given different quarantining strategies. The tests are assumed to all be PCR, and testing starts when 20% of the population has recovered. (a) Enforcing the intervention when the individuals are sampled leads to a higher peak, however the peak remains between 4% to 6% of the population. (b) This translates to around 15% to 25% of homes quarantined.

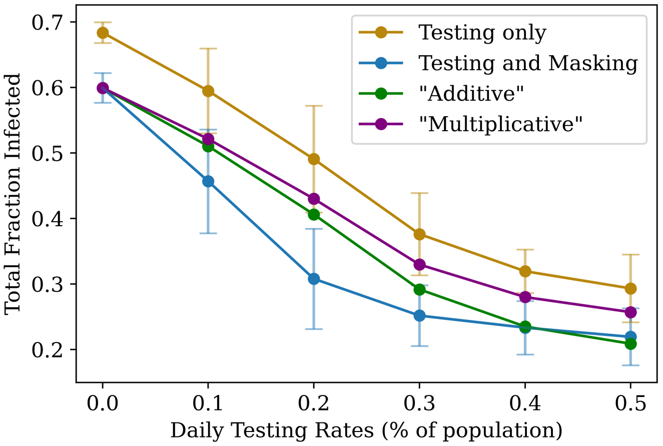


**Fig 6.1: Demonstrating the synergy between testing and masking.** The effects of masking on testing is found to be lower than if it had been purely additive or multiplicative.

tiplicative. For a given daily testing rate of $r$, let the function $f(r)$ describe the effect of testing (without masking), and $g(r)$ describe the effect of testing and masking combined. We further define $\Delta N = f(0) - g(0)$ to be the effect of purely masking the population. Then, by an additive effect we mean that:

$$h_{\text{add}}(r) = f(r) - \Delta N,$$

and by a multiplicative effect we mean that

$$h_{\text{mult}}(r) = f(r) \times \left( \frac{g(0)}{f(0)} \right).$$

All four functions $f(r), g(r), h_{\text{add}}(r)$, and $h_{\text{mult}}(r)$ are shown in Fig. 6.1

#### S7 Flowcharts of Simulation

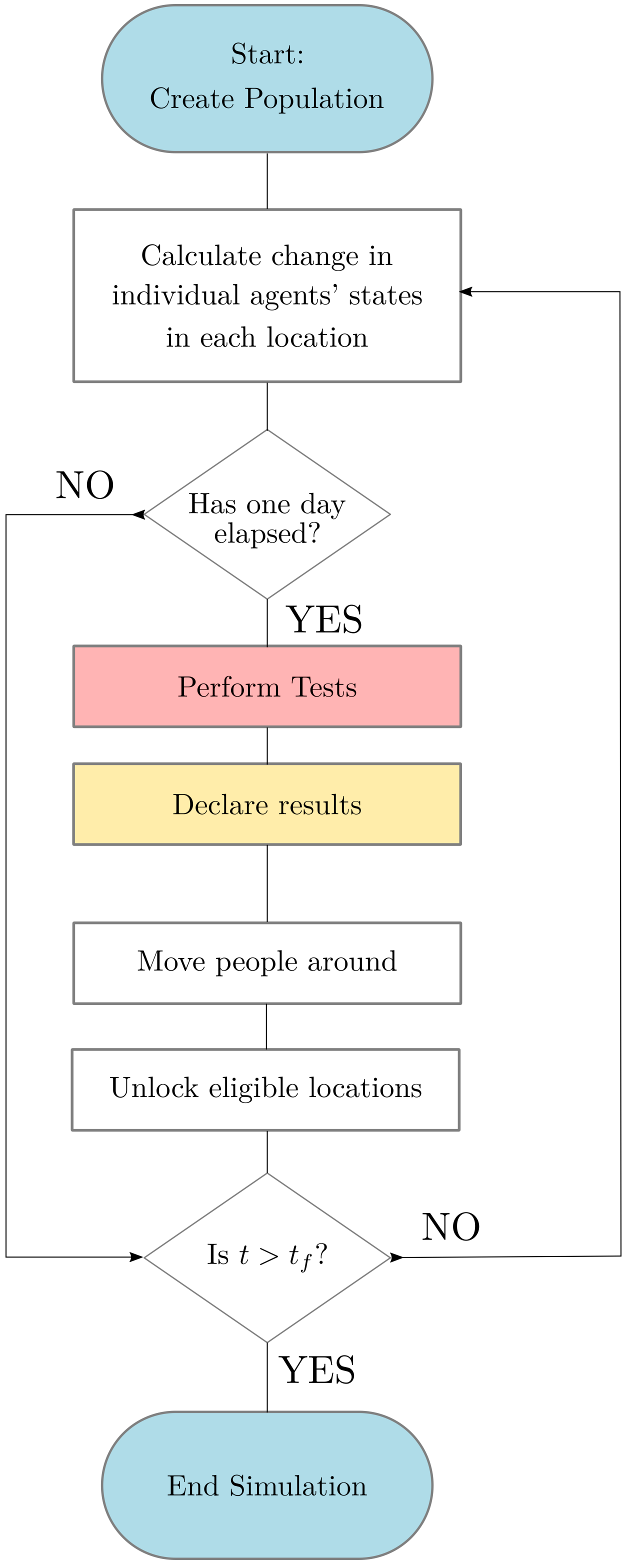


**Fig 7.1:** Global schematic of the simulation.

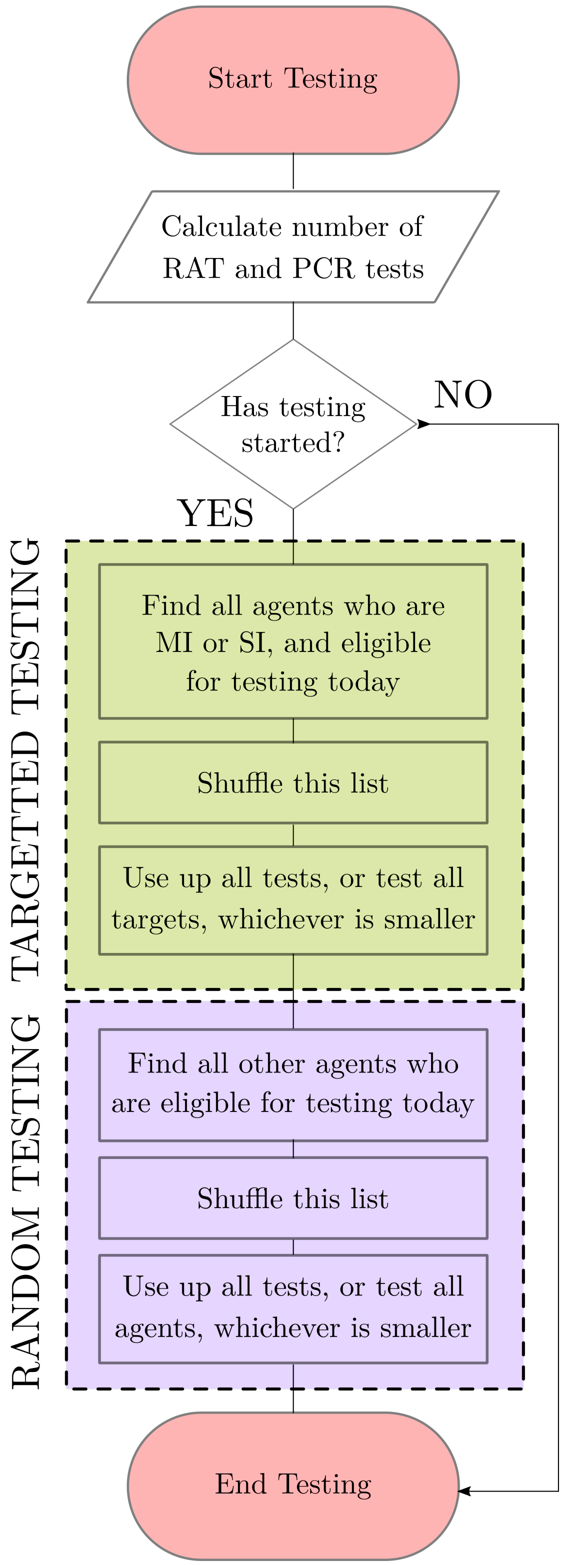


**Fig 7.2:** Schematic of testing individuals.

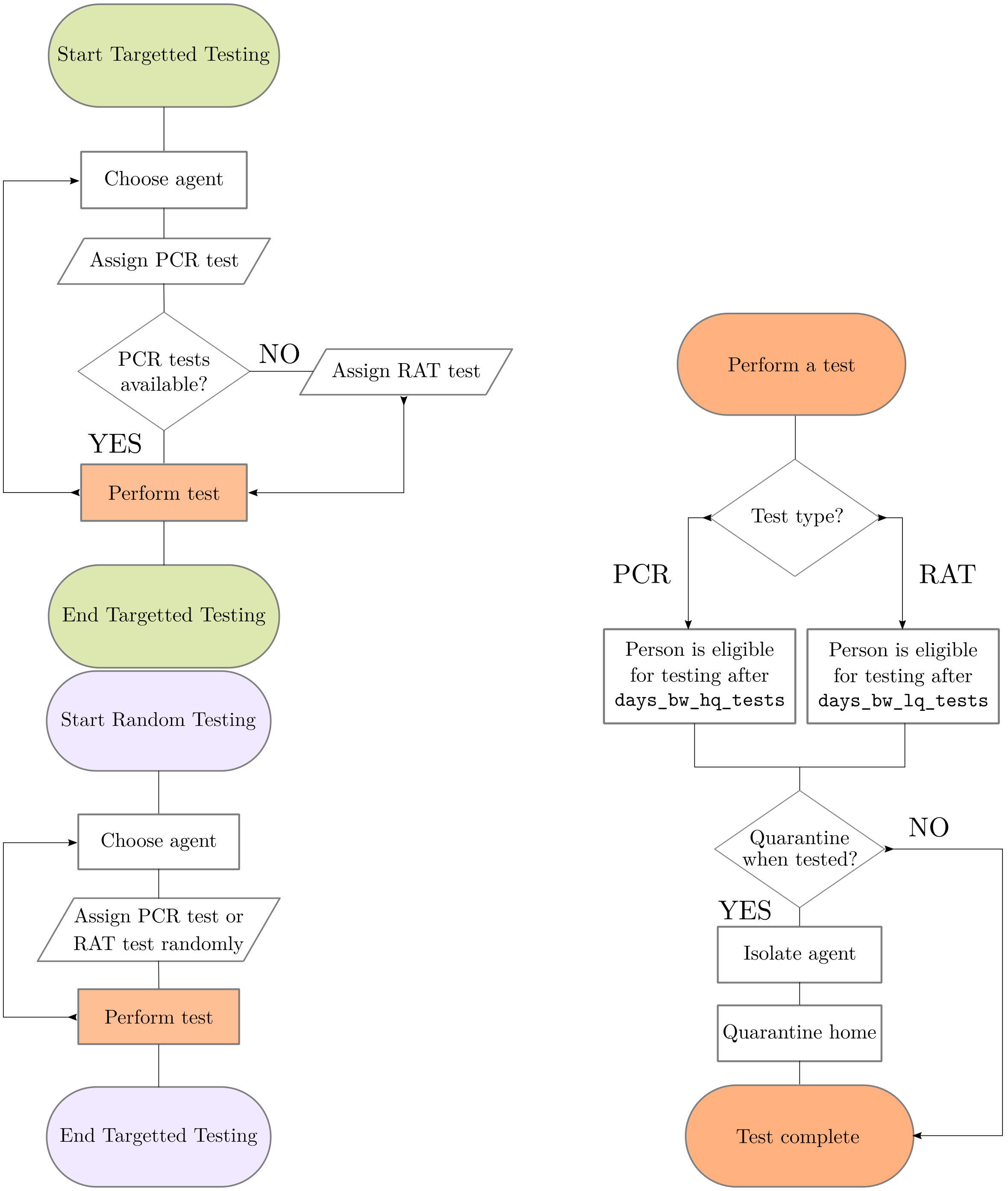


**(a)** Schematic of targeted and random testing.

**(b)** Schematic of performing a test.

**Fig 7.3:** Schematic of testing procedures.

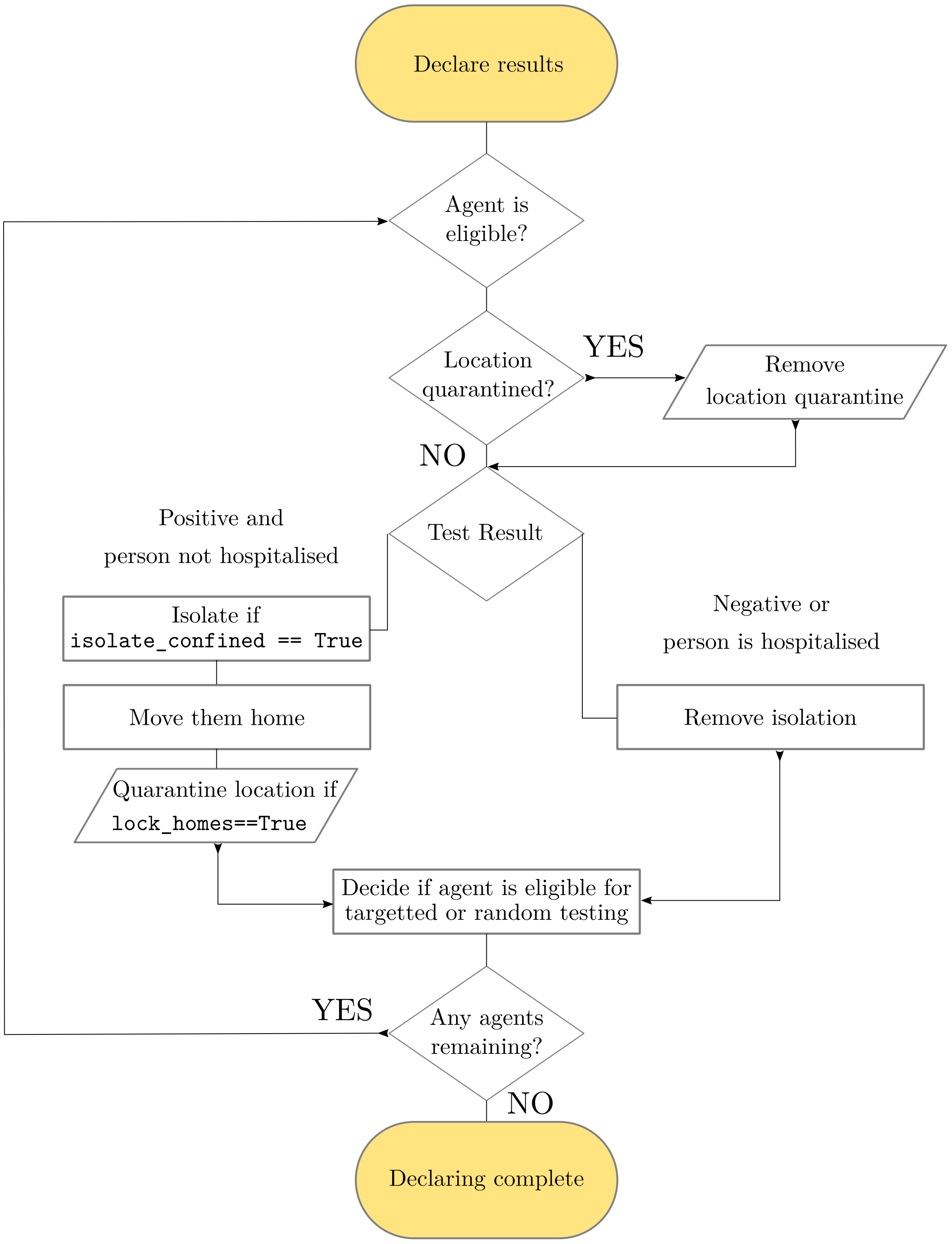


**Fig 7.4:** Schematic of declaring results.
